# Greater than the sum of its parts: A novel psychiatric multimorbidity metric applied to rare disease

**DOI:** 10.64898/2026.11.05.26367370

**Authors:** Rebecca M Pollak, Esra Sefik, Cheryl Klaiman, Daniela V Bishop, Matthew K Harner, Ariana Khan, John R Purcell, Madeleine Wheeler, Teresa Irving, Stormi Pulver, Joseph F Cubells, Elaine F Walker, Melissa M Murphy, Jennifer Gladys Mulle

**Author notes:** Corresponding author information: Rebecca M Pollak, PhD; Center for Advanced Biotechnology and Medicine and Department of Psychiatry, Robert Wood Johnson Medical School, Rutgers University, 679 Hoes Lane West, Piscataway, NJ 08854.

## Abstract

**Importance:** Rare genetic syndromes are often drivers of complex multimorbidity. However, current methods consider multimorbidity as the sum of chronic diagnoses, which does not account for subthreshold impairment.

**Objective:** To develop a quantitative multimorbidity score (MMS) for rare genetic syndromes that summarizes the neurodevelopmental and psychiatric multimorbidity landscape, including subthreshold symptoms, using 3q29 deletion and 3q29 duplication syndromes as hallmark examples.

**Design:** We developed a multimorbidity metric and applied it to cross-sectional data. Remote phenotyping from 2023-2026. In-person phenotyping from 2017-2020.

**Setting:** Remote phenotyping conducted at Rutgers University. In-person phenotyping conducted at Emory University.

**Participants:** Participants were 6 years of age and older. 3q29del and 3q29dup recruited from the 3q29 registry and direct referrals and contacts. Syndrome status confirmed by genetic report review. Typically developing controls recruited from community sources and ResearchMatch.

**Main Outcomes and Measures:** We developed a quantitative MMS across five distinct domains of functioning, operationalized as the sum of z-scores across six standardized assessments. The MMS was calculated for each study participant and compared across groups. Within 3q29del, we evaluated the relationship between MMSs and Vineland-3 adaptive behavior.

**Results:** 96 individuals with 3q29del (52.08% male, 18.01±11.47 years old), 28 individuals with 3q29dup (50.00% male, 20.82±13.64 years old), and 53 controls (41.51% male, 17.77±10.74 years old) were included. 3q29del and 3q29dup participants had significantly lower MMS scores compared to controls (3q29del mean=-8.22±4.67, 3q29dup mean=-6.47±4.66, control mean=1.36±2.35, 3q29del p<2E-16, 3q29dup p=4.89E-16). Within 3q29del, subjects with more severe multimorbidity had greater impairment in adaptive behavior (R^2^=0.589, p<2E-16). The quantitative MMS is more highly correlated with adaptive behavior than traditional diagnosis-based multimorbidity metrics (quantitative MMS R^2^=0.589, diagnosis-based multimorbidity R^2^=0.447). In a subset of 24 3q29del participants with longitudinal data (58.33% male, 13.17±9.08 years old at first evaluation, average time to follow up=5.17±0.87 years), the MMS is stable over time (R^2^=0.858, p=8.34E-8).

**Conclusions and Relevance:** Our novel quantitative MMS is an improved metric for measuring psychiatric multimorbidity in high-risk populations, and accounts for the impact of subthreshold impairments. When applied to 3q29del and 3q29dup, it quantifies the individual burden of multimorbidity, is stable over time, and is associated with clinical outcome measures.

**Key Points:** *Question:* In rare genetic syndromes, can we develop a quantitative, holistic, cross-dimensional metric of neurodevelopmental and psychiatric multimorbidity?

*Findings:* We developed a novel quantitative multimorbidity score that synthesizes data across five phenotypic domains and applied it to cross-sectional data from 96 participants with 3q29 deletion syndrome, 28 participants with 3q29 duplication syndrome, and 53 neurotypical controls. More severe psychiatric multimorbidity was significantly associated with worse adaptive behavior, and quantitative and discretized versions of our score were more informative than counting diagnoses.

*Meaning:* Our novel quantitative multimorbidity score is more informative than traditional metrics and incorporates the impact of subthreshold symptoms, rendering a more complete picture of an individual’s level of impairment.

## Introduction

There is a growing appreciation in the clinical literature for the concept of multimorbidity, defined as the presence of multiple chronic diseases in a single individual^1,2^. Individuals with multimorbidity have worse outcomes than would be expected based on the additive effects of their individual diagnoses^3,4^. Many multimorbidity studies include both physical and psychiatric diagnoses and have largely been conducted on older adult populations^5–7^. Psychiatric multimorbidity focuses exclusively on neurodevelopmental and psychiatric diagnoses; studies in the general population have shown elevated rates of psychiatric multimorbidity among adults with severe mental illness^8,9^, including schizophrenia (SZ)^10^. Psychiatric disorders diagnosed in adolescence are also strong predictors of later psychiatric multimorbidity^11^. Neurodevelopmental disorders, especially autism spectrum disorder (ASD), intellectual disability (ID), and attention deficit/hyperactivity disorder (ADHD), are also associated with increased risk for psychiatric multimorbidity in adults^12^, and co-occurrence of these disorders is associated with worse school performance in children^13^.

Copy number variant (CNV) disorders are associated with increased risk for an array of medical and psychiatric diagnoses. Among patients with a CNV, multimorbidity is common. For example, the rate of general multimorbidity in young adults with 22q11.2 deletion syndrome is markedly higher than rates in age-matched controls^14,15^, and is similar to multimorbidity rates in substantially older general population samples^16^. Individuals with 22q11.2 deletion syndrome, 16p11.2 deletion syndrome, 16p11.2 duplication syndrome, and 22q11.2 duplication syndrome also experience elevated rates of <u>psychiatric</u> multimorbidity^17,18,19^. However, multimorbidity models have typically relied on binary, present/absent categorization of diseases^5,20,21^, and thus do not account for subthreshold impairment that may contribute to poor outcomes. Subthreshold phenotypes are well-documented in 22q11.2 deletion syndrome and other rare genetic syndromes^18,22,23^, but these are not captured by existing multimorbidity approaches.

3q29 deletion syndrome (3q29del) and 3q29 duplication syndrome (3q29dup) are rare (∼1:30,000) CNV disorders characterized by a 1.6 Mb deletion or duplication on the long arm of chromosome 3 (hg19, chr3:195725000-197350000)^24–26^. 3q29del is similar to 22q11.2 deletion syndrome and 16p11.2 duplication syndrome, with elevated risk for ASD, ID, ADHD, SZ, and anxiety, as well as multiple medical diagnoses^23–40^. The phenotypic spectrum of 3q29dup is less well-defined; it is associated with increased risk for ASD, ID, and seizures, as well as medical diagnoses^24,41,42^. Interestingly, and unlike 22q11.2 deletion syndrome, somatic phenotypes in 3q29del and 3q29dup are relatively mild; most of the disability burden is due to neurodevelopmental and psychiatric manifestations. Previous work by our team has shown that within subjects with 3q29del, an increasing number of psychiatric diagnoses is associated with impaired adaptive ability and executive function^29,43^. However, psychiatric multimorbidity has not been systematically assessed in this population. Most importantly, this prior work has not considered whether individual patient outcomes are influenced by the cumulative impact of subthreshold impairment across domains.

In the present study, we developed a novel, fully quantitative measure of psychiatric multimorbidity that incorporates data from six distinct standardized assessments across five neurodevelopmental and psychiatric domains; we then applied that measure to 3q29del and 3q29dup. We interrogated the relationship between our novel quantitative multimorbidity score and adaptive ability, and compared our quantitative multimorbidity score to more traditional, count-based models. Finally, we assessed the longitudinal stability of our multimorbidity score in a subset of subjects with 3q29del. This quantitative multimorbidity score may more accurately captures psychiatric multimorbidity in high-risk CNV disorders.

## Methods

### Study participants

Individuals with 3q29del (n=96, 52.08% male, 18.01±11.47 years old) and 3q29dup (n=28, 50.00% male, 20.82±13.64 years old) were recruited for a deep phenotyping study that was conducted remotely^44^. Participants were recruited from the existing online 3q29 registry (3q29deletion.com)^25^, as well as direct referrals and contacts. Individuals with a 3q29 deletion or duplication residing completely outside of the canonical 1.6 Mb interval were excluded from analysis. Typically developing controls (n=53, 41.51% male, 17.77±10.74 years old) were recruited from community sources and from ResearchMatch, an online platform for individuals interested in participating in research^45^.

### Measures

Prior deep phenotyping of individuals with 3q29del has revealed the following major axes of disability: cognition, social function, psychosis, anxiety, executive function and attention, and visual-motor integration^27,28^. The measures used in this study were as previously described (Supplemental Material)^44^. A subset of 3q29del participants (n=24, 58.33% male, 13.17±9.08 years old at first evaluation) had been previously directly evaluated by our team^27,28,46^ (Table S1).

### Multimorbidity score calculation

The multimorbidity score (MMS) was developed as follows: scores on the standardized assessments were each converted to z-scores based on each instrument’s normative mean and standard deviation. Scales were aligned such that impairment on any measure corresponds to a negative z-score. The composite MMS is the sum of the z-scores across all assessments, where a more negative MMS corresponds to more severe impairment. The MMS was calculated for each participant with complete data across all six assessments.

### Analysis

All analyses were performed in R version 4.2.2^47^. All regressions were adjusted for age and sex. Statistical analysis was performed using simple linear models implemented using the stats R package^47^. Model fit was compared using Cox tests implemented using the lmtest R package^48^. Data visualization was performed using the ggplot2^49^ and corrplot^50^ R packages. A leave-one-out analysis showed that the CBCL/ABCL DSM-keyed anxiety problems score did not meaningfully improve the association between the MMS and the Vineland ABC score (Table S2). However, we included the CBCL/ABCL DSM-keyed anxiety problems score in the MMS calculation because of the substantial burden of anxiety phenotypes in 3q29del^27^.

## Results

### Multimorbidity burden in 3q29del and 3q29dup

Participants with 3q29del had significantly lower MMSs than typically developing controls (3q29del mean=-8.22±4.67, range -21.67-3.38; control mean=1.36±2.35, range -4.91-8.44; p<2E-16; Figure 1A). Participants with 3q29dup also had significantly lower MMSs than controls (3q29dup mean=-6.47±4.66, range -13.59-2.43; p=4.89E-16; Figure 1A). The multimorbidity burden in 3q29dup is less than that in 3q29del; however, the difference does not reach statistical significance (p=0.164; Figure 1A) likely due to the small sample size for 3q29dup. Across all subjects, we found a strong positive correlation between the total MMS and the Vineland Adaptive Behavior Composite (ABC) (adjusted R^2^=0.719, p<2E-16; Figure 1B), where individuals with lower MMSs have greater impairments to adaptive ability. No single MMS component assessment replicated this correlation (Figure 1C), demonstrating that the total MMS is the best predictor of adaptive ability. Using Pearson correlation analysis, we found different relationships among the six MMS components based on genotype, suggesting there are genotype-specific differences in the relationship between MMS component scores (Supplemental Material).

**Figure 1.**
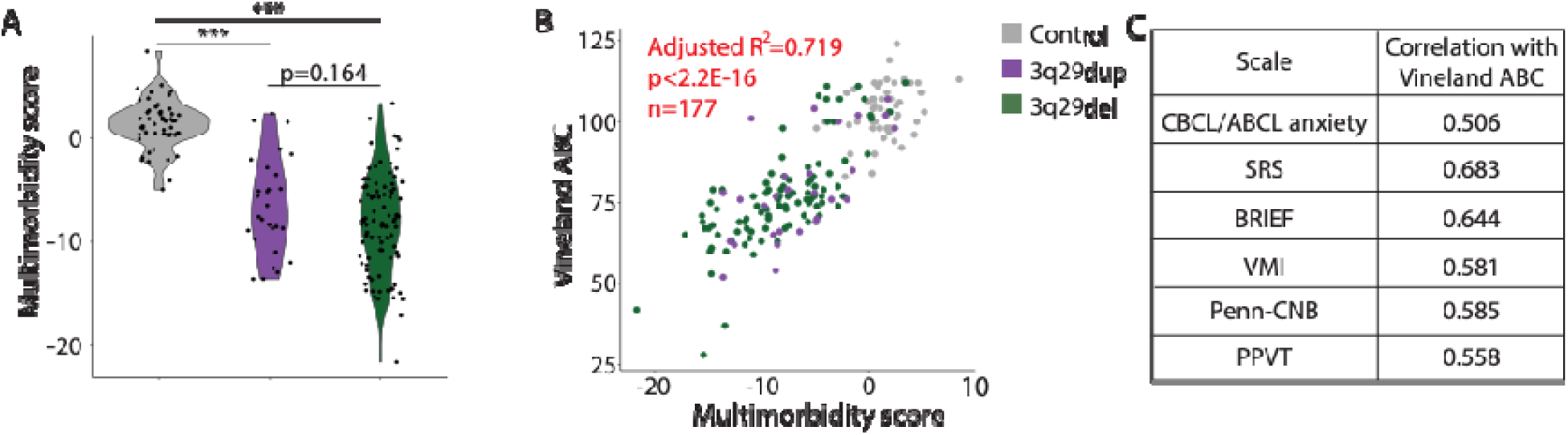
**A)** Distribution of multimorbidity scores for control (n=53), 3q29del (n=96), and 3q29dup (n=28) study participants. **B)** Correlation between total multimorbidity score and Vineland ABC score for control, 3q29del, and 3q29dup participants. **C)** Table showing the Pearson correlation of individual multimorbidity score components with the Vineland ABC. ***, p<0.001

To further understand the MMS composition, we visually examined the contributions of the six assessments to the total MMS (Figure 2). We did not observe a specific domain or subset of domains were driving the total MMSs in 3q29dup and 3q29del; rather, all tasks contribute to the score, with no one task or domain having an outsize impact (Figure 2B-C). These data demonstrate a substantial degree of multimorbidity in 3q29dup and 3q29del relative to typically developing controls and show that all six tasks included in our MMS contribute to total scores.

**Figure 2.**
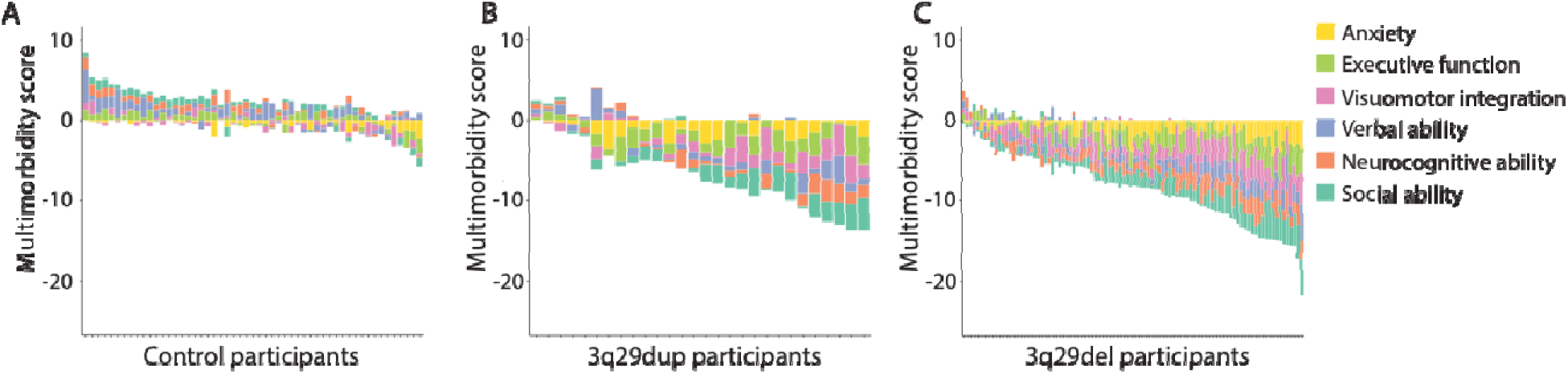
**A)** Contribution of each multimorbidity score item to total multimorbidity scores for control participants (n=53). **B)** Contribution of each multimorbidity score item to total multimorbidity scores for 3q29dup participants (n=28). **C)** Contribution of each multimorbidity score item to total multimorbidity scores for 3q29del participants (n=96).

### The MMS outperforms traditional multimorbidity metrics for 3q29del

In the current literature, multimorbidity is typically quantified as the arithmetic or weighted sum of clinical diagnoses^5,20,21^. Within the 3q29del sample, we tested whether the quantitative MMS is a better descriptor of disability in our sample by comparing it to an approximation of traditional methods. We defined a “putative diagnosis” in our sample as any item with a score more than two standard deviations from the normative population mean (range=0-6 putative diagnoses, Table S3). The number of putative diagnoses had a moderate correlation with the Vineland ABC score (adjusted R^2^=0.447, Figure S3A) but did not fully replicate the association between the quantitative MMS and the Vineland ABC in 3q29del (adjusted R^2^=0.589) (Figure S3D, Table S4). It is possible that the MMS-Vineland association is aided by the quantitative nature of the MMS, which is a function of statistical power but not necessarily clinically meaningful. To test this, we discretized the quantitative MMS distribution using two additional methods. (Table S3). First, we divided the MMS distribution into seven groups, with the same number of individuals per group (n=13-14 subjects per group). Second, we divided the distribution into seven equal-width bins, with varying numbers of individuals per group (n=1-31 subjects per group). The MMS divided into equal groups or equal bins did not fully replicate the association between the quantitative MMS and the Vineland ABC score (equal groups adjusted R^2^=0.529, equal bins adjusted R^2^=0.518, Figure S3B-C, Table S4). Notably, both discretized versions of the MMS distribution had stronger associations with the Vineland ABC score than the putative diagnoses model, demonstrating that even our discretized MMS is more informative than traditional multimorbidity metrics.

To illustrate the resolution of the MMS, we provide three examples from within our 3q29 deletion sample. Figure 3A shows metrics for two individuals who each have zero putative diagnoses; by traditional multimorbidity metrics, these individuals would both fall into the same category of “minimally affected.” However, their adaptive behavior, as measured by their Vineland ABC scores, are quite different (71 vs 112) and this is represented by their MMS (-6.04 vs 3.38). This is due to the consistently low but sub-diagnostic threshold performance of individual #1 across all domains, which likely has a cumulative impact on functioning (Figure 3B). Similarly, individuals 3 and 4 (Figure 3C-D) both have two putative diagnoses, yet have Vineland ABC scores of 37 and 98, respectively. Individual #3, with a Vineland ABC score of 37, has consistently low functioning across domains. Individual #4, with a Vineland ABC score of 98, displays higher function in several domains but has significant anxiety and compromised executive function. Their overall phenotype is well-described by the MMS (-13.43 vs -8.03). These data demonstrate that simply counting diagnoses does not provide the same resolution as our quantitative MMS, and that the degree of disability can vary widely even between individuals with the same number of putative diagnoses.

**Figure 3.**
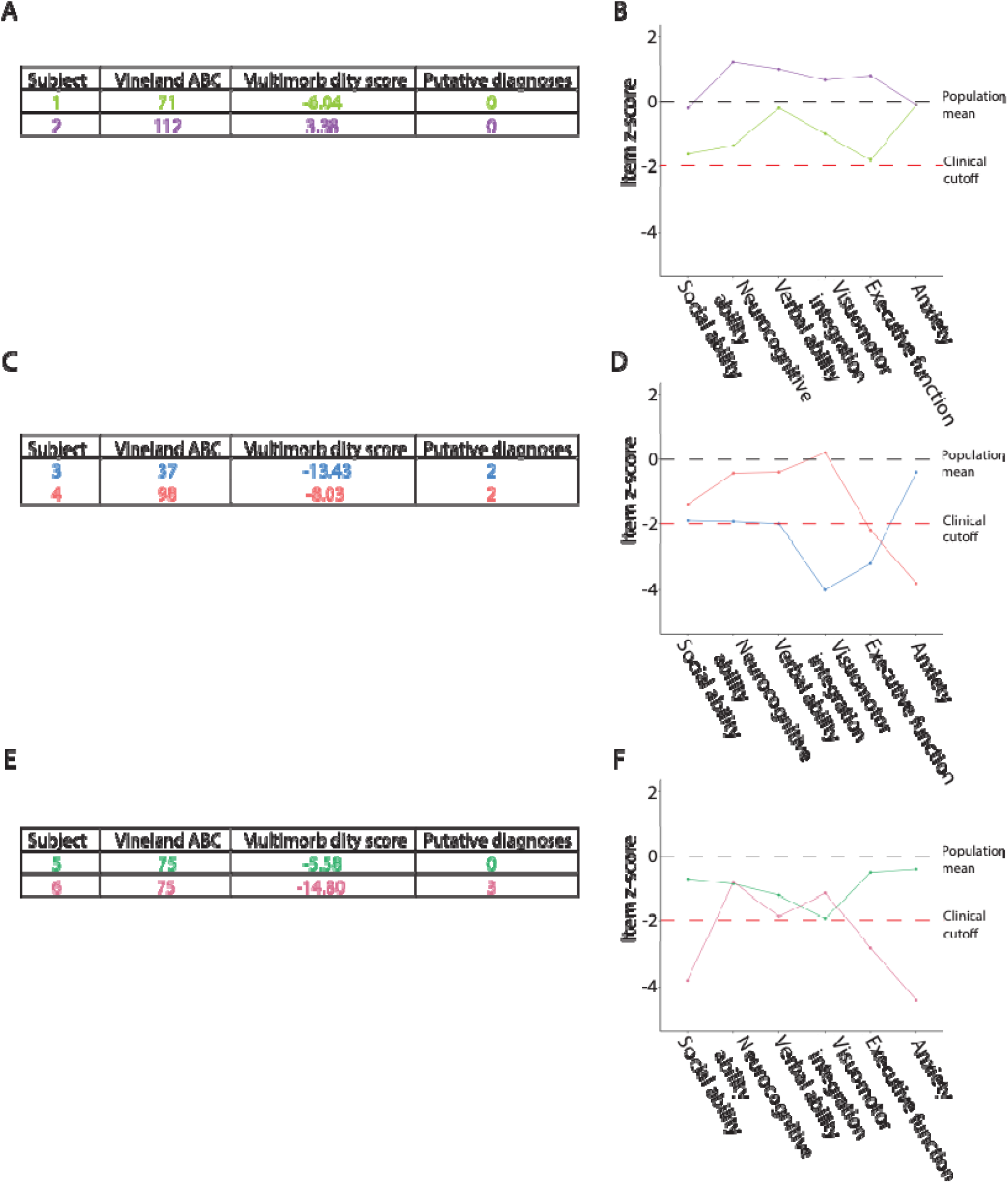
**A)** Summary of exemplar 3q29del participants with no putative diagnoses. **B)** Profile of z-scores for each multimorbidity score item for exemplar 3q29del participants with no putative diagnoses. **C)** Summary of exemplar 3q29del participants with two putative diagnoses. **D)** Profile of z-scores for each multimorbidity score item for exemplar 3q29del participants with two putative diagnoses. **E)** Summary of exemplar 3q29del participants with a Vineland ABC score of 75. **F)** Profile of z-scores for each multimorbidity score item for exemplar 3q29del participants with a Vineland ABC score of 75.

Is the MMS redundant with the Vineland? To answer this question, we compared the MMSs of individuals with the same Vineland ABC score. Figure 3E-F shows two individuals with a Vineland ABC score of 75; however, individual #5 has a MMS of -5.58 and no putative diagnoses, while individual #6 with the same Vineland ABC score has a MMS of -14.80 and three putative diagnoses. These individuals would be well-described by traditional multimorbidity metrics that count diagnoses, but their differences would not be captured by the Vineland. Thus while the quantitative MMS and the Vineland ABC score are highly correlated in 3q29del, the MMS gives a more informative picture of the scope of disability for a given individual. Taken together, these data highlight how the MMS is an effective compromise between single outcome measures (such as adaptive behavior as measured by the Vineland) and traditional multimorbidity measures that rely on counts of diagnoses within an individual.

### Multimorbidity in 3q29del is stable over time

Our team has previously evaluated a subset of our study participants (n=24, 58.33% male, Table S2) as part of an in-person deep phenotyping study^27,28,46^. To evaluate the stability of our quantitative MMS over time, we compared the MMS from their original evaluation scores to the MMS from the present study (average time to follow up=5.17±0.87 years). The MMSs were remarkably stable over time, with a strong positive correlation between timepoints (R=0.827, p=6.38E-7; Figure 4A-B). We also found that the contributions of the different score items remained relatively stable from the first evaluation to the second (Figure 4C-D). Notably, the scores remained stable across time, assessment format (in-person versus virtual), and cognitive assessment tools (DAS-II/WASI-II^27,28,46^ versus Penn-CNB/PPVT^44^). This further emphasizes the stability of our quantitative MMS and suggests that our score is capturing disability related to core traits, rather than state-based phenotypes.

**Figure 4.**
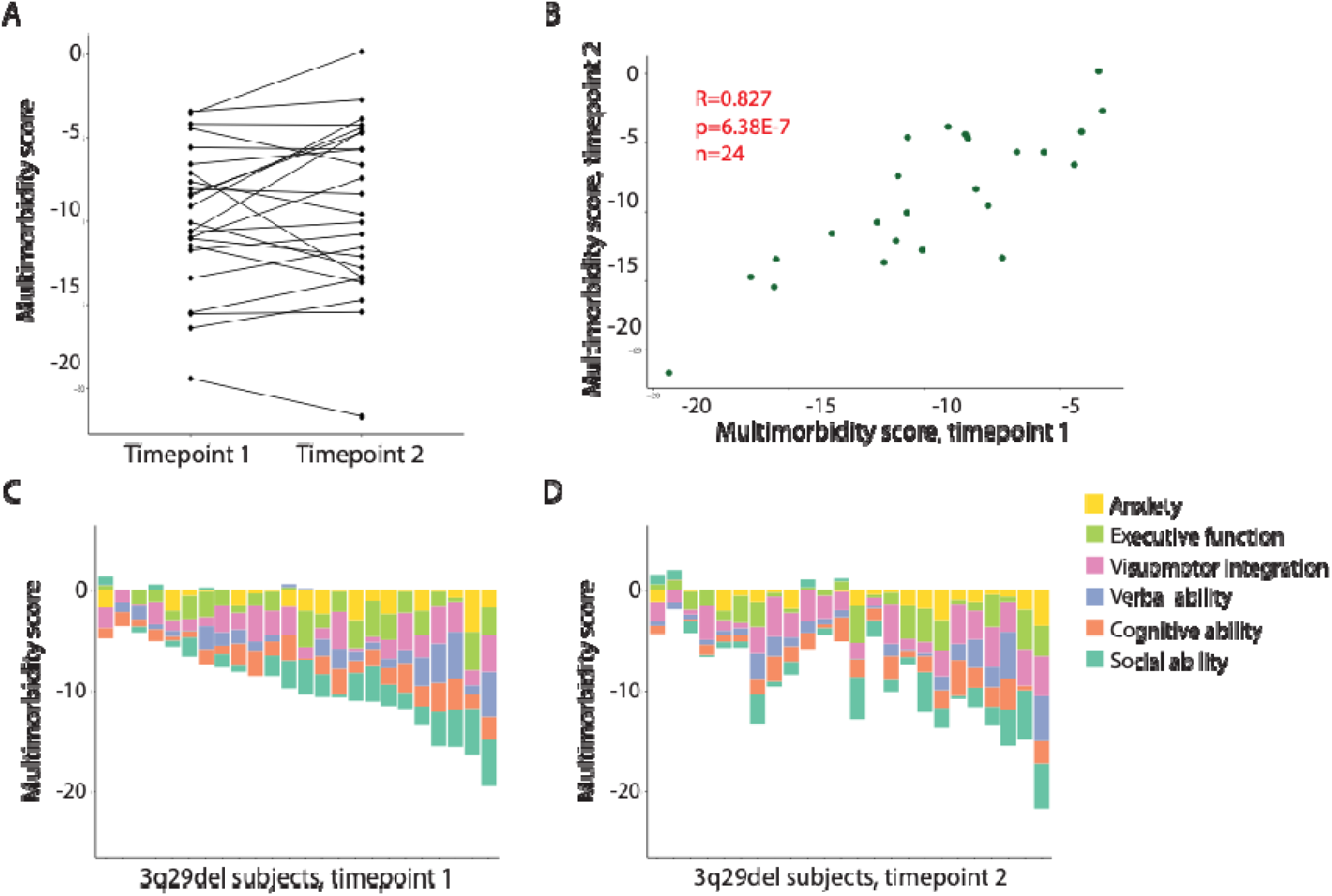
**A)** Line plot showing the change in total multimorbidity score between evaluations for 3q29del participants with longitudinal data (n=24). **B)** Correlation between the total multimorbidity score at timepoint 1 and the total multimorbidity score at timepoint 2 for 3q29del subjects with longitudinal data (n=24). **C)** Contribution of each multimorbidity score item to total multimorbidity scores for longitudinally assessed 3q29del participants at timepoint 1. **D)** Contribution of each multimorbidity score item to total multimorbidity scores for longitudinally assessed 3q29del participants at timepoint 2. Note that the order of subjects in panels C and D is identical.

## Discussion

The present study is the first to introduce a novel, fully quantitative measure of psychiatric multimorbidity that can account for subthreshold impairment, applied to two rare CNV disorders. We found that subjects with 3q29del and 3q29dup have a significantly increased burden of multimorbidity as compared to typically developing controls, and increased multimorbidity was associated with greater impairments to adaptive ability as measured by the Vineland. Compared to traditional measures, the MMS we developed revealed more nuanced information into the total burden of multimorbidity as well as the specific behavioral domains with the most deficits. The quantitative MMS was also more informative than examining the Vineland ABC score alone, highlighting the value of the composite score in assessing the full scope of multimorbidity associated with 3q29del. Finally, we found that multimorbidity burden is relatively stable over time in a subset of 3q29del participants with longitudinal data.

In the present study we focused specifically on psychiatric multimorbidity, as individuals with 3q29del are at substantially increased risk for a wide range of neurodevelopmental and psychiatric disorders^27^. Recent studies of psychiatric multimorbidity in the phenotypically similar 16p11.2 deletion and duplication and 22q11.2 deletion and duplication syndromes have identified an increased burden of psychiatric multimorbidity in affected individuals^17,18^. While these studies did not create a composite quantitative multimorbidity metric, they did find that a majority of individuals with 16p11.2 and 22q11.2 CNVs have clinically significant or subthreshold impairment across multiple clinical domains^17,18^, consistent with our findings in 3q29del. Additionally, as the number of affected phenotypic domains in 16p11.2 and 22q11.2 CNV carriers increases, the average Vineland ABC score decreases^17,18^, again consistent with the results of the present study in 3q29del. Together, these data highlight that increased psychiatric multimorbidity is a common feature among CNV disorders, and suggest that our MMS could be applied across disorders to more fully dissect the impact of both clinically significant and subthreshold impairment across domains.

Most studies of multimorbidity in the literature are based on the number of chronic clinical diagnoses an individual has received, typically via medical records review^5,20,21^. Some multimorbidity metrics attempt to account for the fact that some clinical diagnoses have a greater impact on quality of life and long-term outcomes compared to other diagnoses by weighting the different diagnoses in the calculation of the score^20,21^. However, diagnosis-based models do not capture the cumulative impact of subthreshold impairment across multiple domains, and do not account for subthreshold impairment in some domains possibly modulating the clinical presentation of diagnoses in other domains. We tested four different variations of our MMS to assess whether incorporating truly quantitative measures improves the performance of the score. Across all comparisons, we found that the quantitative version of our MMS is the most informative; additionally, discretized versions of our MMS were still more informative than a count of affected domains. Our MMS was also more informative than the Vineland ABC alone, demonstrating that our quantitative MMS gives a more informative summary of impairment across multiple clinically relevant domains.

Phenotypic heterogeneity in genetic disorders introduces challenges for the choice of clinical trial endpoints. With the advent of precision medicine initiatives and emerging advances in genetic therapies, the selection of appropriate outcome measures for clinical trials takes on new urgency. For example, Fragile X syndrome has been the subject of intense drug development efforts, with over 20 clinical trials to date that have met with little success^51,52^. While the reasons for this are manifold, it has been suggested that phenotypic heterogeneity among patients, and the inability of outcome measures to account for this heterogeneity, may be a casual factor^53^. It has been further suggested that “aggregate measures composed of several individual outcome measures” may represent an avenue to better capture treatment effects^52^. Given that the MMS described here is composed of functional measures across domains, incorporates both subjective caregiver-derived evaluations and objective task-based performance, and is stable over time, it may be informative as a possible clinical trial endpoint in genetic disorders with high psychiatric multimorbidity. This holistic, cross-domain approach may serve as an informative measure of treatment efficacy, representing a possible use case for the MMS we have developed here.

While this is the first study to comprehensively assess psychiatric multimorbidity in 3q29del and 3q29dup, it is not without limitations. First, the measures included in the MMS calculation are largely parent- or caregiver-report screening tools rather than diagnostic instruments, limiting our ability to assess true clinical impairment across domains. However, using remotely deployed screening tools increases the feasibility of incorporating the calculation of a composite MMS into standard of care practices for high-risk individuals. This may enable care providers to identify more severely impacted individuals at earlier timepoints, before severe clinical impairment is evident, allowing for deployment of early intervention strategies. Second, the CBCL/ABCL DSM-keyed anxiety problems score does not contribute to the MMS’s correlation with Vineland ABC scores; we kept this measure in the MMS calculation because anecdotal evidence shows that anxiety has significant impact on quality of life for individuals with

3q29del. However, this instrument may not be fully capturing the impact of anxiety in our study population, and a different measure may better quantify its effects. Third, we were unable in the present study to assess the influence of race and ethnicity on the expression of multimorbidity in individuals with 3q29del and 3q29dup. Future recruitment efforts targeting underserved populations will help to address this question. Finally, the sample size for our longitudinal analysis was small, limiting the predictive power of the analysis. Future longitudinal studies will allow us to more comprehensively explore the landscape of psychiatric multimorbidity in 3q29del over time.

This is the first description of a novel quantitative multimorbidity score, and the first application of this metric to describe psychiatric multimorbidity in 3q29del and 3q29dup. We identified a significant burden of psychiatric multimorbidity in 3q29del and 3q29dup, and subjects with more severe multimorbidity had worse adaptive ability as measured by the Vineland. Our multimorbidity score was more informative than traditional multimorbidity metrics and revealed the impact of subthreshold impairment on the total multimorbidity burden. In a subset of 3q29del participants with longitudinal data multimorbidity burden was stable over time, suggesting that our novel multimorbidity score is quantifying a stable trait associated with 3q29del. Together, these data emphasize the role of psychiatric multimorbidity in the 3q29del phenotype, and suggest that measuring multimorbidity may improve our prediction of long-term outcomes for affected individuals.

## Data Availability

All data produced in the present study are available upon reasonable request to the authors.

## Data access

Rebecca M Pollak and Jennifer G Mulle had full access to all the data in the study and take responsibility for the integrity of the data and the accuracy of the data analysis.

## Data sharing statement

Data are available from the corresponding author upon reasonable request.

**Table 1.** Demographics of study participants.

|  |  | Control |  | 3q29 deletion |  | 3q29 duplication |  |
|---|---|---|---|---|---|---|---|
| N |  | 53 |  | 96 |  | 28 |  |
|  |  | Mean ± SD | Range | Mean ± SD | Range | Mean ± SD | Range |
| Age (years) |  | 17.77 ± 10.74 | 8 - 62 | 18.01 ± 11.47 | 6 - 68 | 20.82 ± 13.64 | 7 - 48 |
|  |  | N | % | N | % | N | % |
| Sex |  |  |  |  |  |  |  |
|  | Male | 22 | 41.51% | 50 | 52.08% | 14 | 50.00% |
|  | Female | 31 | 58.49% | 46 | 47.92% | 14 | 50.00% |
| Race |  |  |  |  |  |  |  |
|  | White | 44 | 83.02% | 88 | 91.67% | 23 | 82.14% |
|  | Other | 9 | 16.98% | 8 | 8.33% | 5 | 17.86% |
| Ethnicity |  |  |  |  |  |  |  |
|  | Hispanic/Latino | 6 | 11.32% | 8 | 8.33% | 2 | 7.14% |
|  | Not Hispanic/Latino | 47 | 88.68% | 88 | 91.67% | 26 | 92.86% |

## Acknowledgments

We gratefully acknowledge our study population, the 3q29 deletion and 3q29 duplication community, for their participation and commitment to research.

## Author contributions

RMP performed analysis, created figures and tables, and wrote the manuscript. ES, CK, DVB, MKH, AK, JRP, MW, TI, SP, JFC, and EFW evaluated study participants. CK, SP, JFC, EFW, and MMM helped with data interpretation. JGM planned and executed the study and was the principal investigator responsible for study coordination.

