## Supplemental Material for "Greater than the sum of its parts: A novel psychiatric multimorbidity metric applied to rare disease"

**Supplemental Methods**

*Measures*

Briefly, anxiety-related symptoms were assessed using the Achenbach Child Behavior Checklist or Adult Behavior Checklist (CBCL/ABCL) DSM-keyed anxiety problems score^1,2^. Executive function was assessed using the Behavior Rating Inventory of Executive Function, Second Edition (BRIEF-2) for participants 18 years of age and younger or the or Behavior Rating Inventory of Executive Function for Adults (BRIEF-A) for participants over 18 years of age^3,4^. Graphomotor ability was assessed using the Beery-Buktenica Developmental Test of Visual-Motor Integration (VMI)^5^. Receptive verbal comprehension was assessed using the Peabody Picture Vocabulary Test, Fifth Edition (PPVT)^6^. Neurocognitive ability was assessed using the Penn Computerized Neurocognitive Test Battery (Penn-CNB)^7^. Social functioning was assessed using the Social Responsiveness Scale (SRS)^8^. Adaptive behavior was assessed using the Vineland Adaptive Behavior Scales, Third Edition, Comprehensive Parent/Caregiver Form (Vineland-3)^9^. The PPVT and Penn-CNB were administered via a HIPAA-compliant videoconferencing platform (Zoom) by a trained research assistant. The CBCL/ABCL, BRIEF-2/BRIEF-A, SRS, and Vineland were completed on REDCap^10,11^ or on the instrument publishers’ websites by a parent or caregiver of the participant. A trained research assistant instructed the parent or caregiver of the participant in the administration of the VMI. Psychosis spectrum symptoms were assessed in a subset of participants (n=83) using the Structured Interview for Psychosis-Risk Syndromes (SIPS)^12^. The SIPS was administered via a HIPAA-compliant videoconferencing platform (Zoom) by a trained research assistant.

**Supplemental Results**

*Multimorbidity burden in 3q29del and 3q29dup*

Using Pearson correlation analysis, we found different correlations among the six MMS components based on genotype. In controls, there was a positive correlation between the anxiety, social, and executive function domains (Figure S1A). In 3q29dup, there were positive correlations between the social and executive function domains, the visuomotor integration and neurocognitive domains, and the neurocognitive and verbal domains; there was a weak negative correlation between the anxiety domain and the neurocognitive and verbal domains (Figure S1B). In 3q29del, there were positive correlations between the anxiety, social, and executive function domains, and between the visuomotor integration, neurocognitive, and verbal domains (Figure S1C). Together, this shows genotype-specific differences in the relationship between MMS component scores.

*Increased multimorbidity is associated with worse clinical outcomes in 3q29del*

As the 3q29 deletion is the highest known risk factor for SZ^13-18^, we tested whether the total multimorbidity score (MMS) was associated with psychosis spectrum symptoms measured using the SIPS. We found a negative correlation between the total MMS and the SIPS total score (adjusted R^2^=0.223, p=4.16E-6; Figure S2A), showing that subjects with 3q29del and worse multimorbidity endorse more psychosis-spectrum symptoms on the SIPS. There was a weak correlation between the total MMS and the SIPS Positive domain score (adjusted R^2^=0.064, p=0.020; Figure S2B). We have found that individuals with 3q29del experience high rates of ADHD^19^; we included the BRIEF-2/BRIEF-A Global Executive Composite score in the total MMS calculation as a quantitative measure of attentional and executive function deficits relevant to this population. However, the SIPS also includes items that can be conflated with ADHD symptomatology. To determine whether the correlations between the total MMS and the SIPS total and Positive domain scores were due to collinearity between the measures, we performed a series of analyses using subsets of the multimorbidity and SIPS scores. We found that excluding the BRIEF-2/BRIEF-A Global Executive Composite score from the total MMS calculation did not eliminate the correlation between the MMS and the SIPS total score (adjusted R^2^=0.178, p=4.18E-5; Figure S2C); excluding the Disorganization domain from the total SIPS score also did not eliminate the correlation between the MMS and the SIPS (adjusted R^2^=0.217, p=7.55E-6; Figure S2D). However, excluding item P5 (Disorganized Communication) from the SIPS Positive domain score eliminates the correlation between the MMS and the SIPS (all p>0.200; Figure S2E-F). Together, these data show limited to no correlation between the total MMS and psychosis-spectrum symptoms as assessed by the SIPS after accounting for collinearity between the measures.

**Supplemental Discussion**

Prior work by our team has identified a significant association between the number of neurodevelopmental and psychiatric diagnoses and adaptive ability^20^. In the present study, we sought to replicate and expand on this finding in a substantially larger sample size of individuals with 3q29del. We found that our novel quantitative MMS had a strong positive correlation with adaptive ability, where individuals with less severe multimorbidity had significantly better adaptive ability. Individuals with 3q29del are also at high risk for SZ^13-15^; we tested whether our quantitative MMS was associated with both the total score and the positive symptom domain score of the SIPS. After accounting for collinearity between our MMS and items on the SIPS we found no significant correlation between the measures, indicating that the burden of psychiatric multimorbidity may be orthogonal to risk for SZ spectrum phenotypes in this population. We also had longitudinal data on a subset of study participants; when we compared the MMSs from their first assessment versus their second assessment we found the scores to be remarkably stable over time. This suggests that our MMS is quantifying stable traits, rather than reflecting the transient state of the study participant at the time of evaluation and further suggests that our MMS may be useful as a predictor of later clinical outcomes. **Table S1.** Demographics for 3q29del study subjects with longitudinal data.

| N | | 24 | |
| --- | --- | --- | --- |
|  |  | Mean ± SD | Range |
| Age (years), timepoint 1 | | 13.17 ± 9.08 | 4 - 39 |
| Age (years), timepoint 2 | | 18.33 ± 9.09 | 10 - 44 |
| Time to follow up (years) | | 5.17 ± 0.87 | 4 - 7 |
|  |  | N | % |
| Sex | |  |  |
|  | Male | 14 | 58.33% |
|  | Female | 10 | 41.67% |
| Race | |  |  |
|  | White | 22 | 91.67% |
|  | Other | 2 | 8.33% |
| Ethnicity | |  |  |
|  | Hispanic/Latino | 1 | 4.17% |
|  | Not Hispanic/Latino | 23 | 95.83% |

**Table S2.** Leave-one-out analysis results for multimorbidity score components. A statistically significant p value indicates that the full MMS is a better fit than the reduced model.

| Score version | AIC | P value versus full MMS |
| --- | --- | --- |
| Full multimorbidity score | 712.55 | - |
| Multimorbidity score without SRS | 728.90 | 2.87E-7 |
| Multimorbidity score without VMI | 722.18 | 0.0005 |
| Multimorbidity score without BRIEF | 724.54 | 5.33E-5 |
| Multimorbidity score without CBCL/ABCL | 697.13 | 0.073 |
| Multimorbidity score without PPVT | 717.65 | 0.015 |
| Multimorbidity score without Penn-CNB | 717.93 | 0.013 |
| Multimorbidity score without cognitive domain | 729.14 | 8.23E-7 |

**Table S3.** Grouping strategy for three different versions of the multimorbidity score.

| Version | Group | Multimorbidity score range | Vineland score range | N | % |
| --- | --- | --- | --- | --- | --- |
| Clinical scales | 0 | -8.57 - 3.38 | 71 – 112 | 28 | 29.17% |
|  | 1 | -9.57 - -3.31 | 68 – 100 | 25 | 26.04% |
|  | 2 | -13.43 - -5.11 | 37 – 98 | 19 | 19.79% |
|  | 3 | -14.96 - -9.58 | 53 – 79 | 13 | 13.54% |
|  | 4 | -17.15 - -10.97 | 28 – 73 | 8 | 8.33% |
|  | 5 | -15.57 - -14.83 | 60 – 71 | 2 | 2.08% |
|  | 6 | -21.67 | 42 | 1 | 1.04% |
| Equal groups | 1 | -3.68 - 3.38 | 74 – 112 | 14 | 14.58% |
|  | 2 | -5.41 - -3.86 | 71 – 111 | 14 | 14.58% |
|  | 3 | -7.47 - -5.42 | 68 – 86 | 13 | 13.54% |
|  | 4 | -8.54 - -7.55 | 63 – 98 | 14 | 14.58% |
|  | 5 | -10.77 - -8.57 | 59 – 84 | 13 | 13.54% |
|  | 6 | -14.10 - -10.79 | 37 – 79 | 14 | 14.58% |
|  | 7 | -21.67 - -14.19 | 28 – 79 | 14 | 14.58% |
| Equal bins | 1 | -0.12 - 3.38 | 90 – 112 | 5 | 5.21% |
|  | 2 | -3.68 - -0.41 | 74 – 111 | 9 | 9.38% |
|  | 3 | -7.07 - -3.86 | 69 – 111 | 24 | 25.00% |
|  | 4 | -10.79 - -7.42 | 59 – 98 | 31 | 32.29% |
|  | 5 | -14.19 - -10.97 | 37 - 79 | 14 | 14.58% |
|  | 6 | -17.15 - -14.57 | 28 – 79 | 12 | 12.50% |
|  | 7 | -21.67 | 42 | 1 | 1.04% |

**Table S4.** Summary of regression model comparison across versions of the multimorbidity score with Vineland ABC as the outcome variable. A statistically significant p value indicates that the full MMS is a better fit than the reduced model.

| Score version | Adjusted R^2^ | AIC | P value versus quantitative MMS |
| --- | --- | --- | --- |
| Quantitative multimorbidity score | 0.589 | 712.55 | - |
| Sum of scales >2 SD from mean (n=1-28 subjects per group) | 0.447 | 745.56 | 3.65E-13 |
| Quantitative score distribution split into 7 equal groups (n=13-14 subjects per group) | 0.529 | 730.21 | 4.50E-6 |
| Quantitative score distribution split into 7 equal-width bins (n=1-31 subjects per group) | 0.518 | 732.39 | 1.89E-5 |

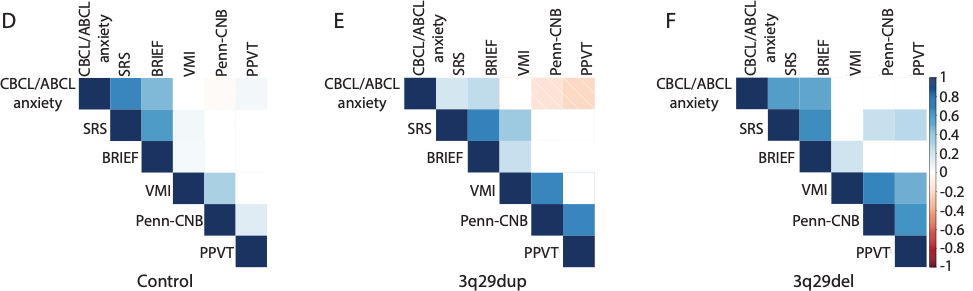

**Figure S1. A)** Pearson correlation matrix showing the relationship between multimorbidity score items in control participants. **B)** Pearson correlation matrix showing the relationship between multimorbidity score items in 3q29dup participants. **C)** Pearson correlation matrix showing the relationship between multimorbidity score items in 3q29del participants.

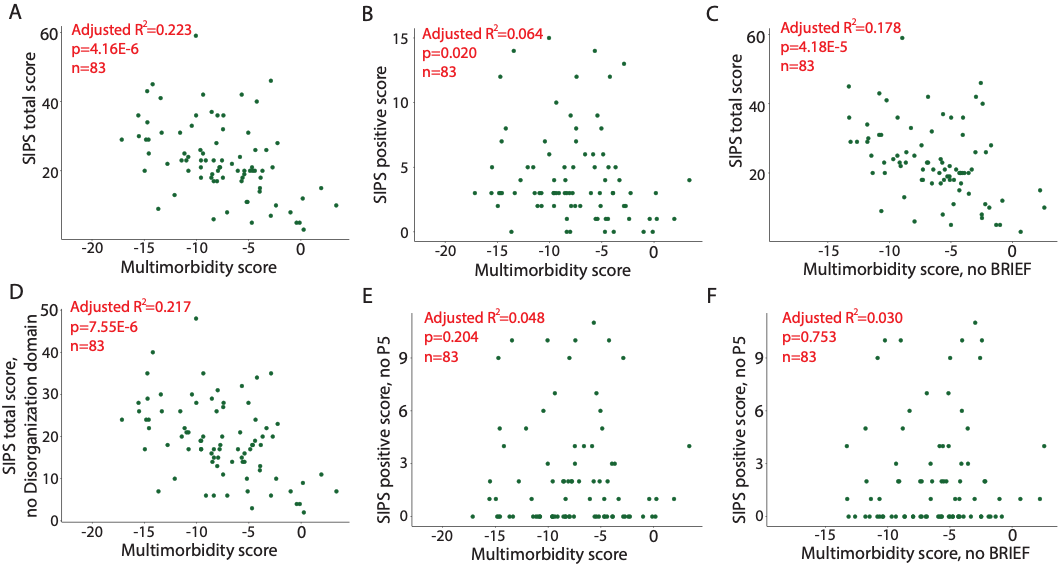

**Figure S2. A)** Correlation between total multimorbidity score and SIPS total score for 3q29del participants (n=83). **B)** Correlation between total multimorbidity score and SIPS Positive domain score for 3q29del participants (n=83). **C)** Correlation between the multimorbidity score, excluding the BRIEF GEC, and the SIPS total score. **D)** Correlation between the multimorbidity score and the SIPS total score, excluding the Disorganization symptom domain. **E)** Correlation between the multimorbidity score and the SIPS Positive domain score, excluding item P5. **F)** Correlation between the multimorbidity score, excluding the BRIEF GEC, and the SIPS Positive domain score, excluding item P5.

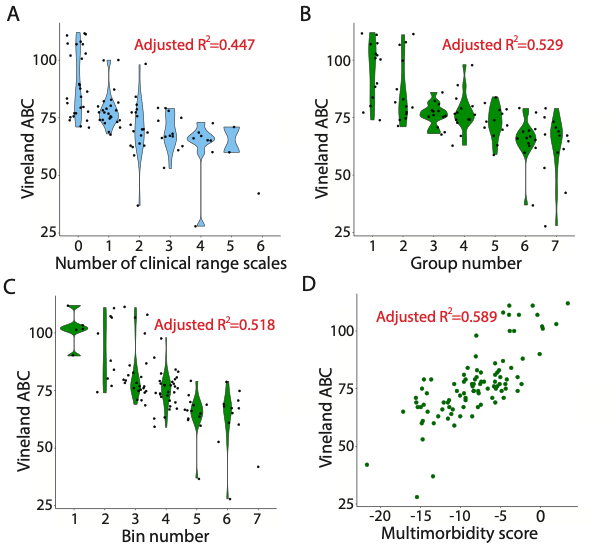

**Figure S3. A)** Distribution of Vineland ABC scores for subjects with 0-6 clinical range scales. **B)** Distribution of Vineland ABC scores for individuals in multimorbidity score groups 1-7, with equal numbers of subjects in each group. **C)** Distribution of Vineland ABC scores for individuals in multimorbidity score bins 1-7, with equal-width bins. **D)** Correlation between total multimorbidity score and Vineland ABC score for 3q29del participants (from Figure 2A).

<https://www.nature.com/articles/mp201440#supplementary-information>

19.	Sanchez Russo R, Gambello MJ, Murphy MM, et al. Deep phenotyping in 3q29 deletion syndrome: recommendations for clinical care. *Genet Med*. May 2021;23(5):872-880. doi:10.1038/s41436-020-01053-1

20.	Pollak R, Burrell T, Cubells J, et al. Adaptive behaviour deficits in individuals with 3q29 deletion syndrome. *Journal of intellectual disability research : JIDR*. Sep 22 2023;doi:10.1111/jir.13094
